# Transcription Factor RUNX1 Regulates Coagulation Factor XIII-A (*F13A1*): Decreased Platelet-Megakaryocyte *F13A1* Expression and Clot Contraction in *RUNX1* Haplodeficiency

**DOI:** 10.1101/2024.12.17.24318561

**Authors:** Fabiola Del Carpio-Cano, Natthapol Songdej, Liying Guan, Guangfen Mao, Lawrence E. Goldfinger, Jeremy G.T. Wurtzel, Kiwon Lee, Michele P Lambert, Mortimer Poncz, A. Koneti Rao

## Abstract

**Background:** Germline *RUNX1* haplodeficiency (RHD) is associated with thrombocytopenia, platelet dysfunction and predisposition to myeloid malignancies. Platelet expression profiling of a RHD patient showed decreased *F13A1,* encoding for the A subunit of factor XIII, a transglutaminase that cross-links fibrin and induces clot stabilization. FXIII-A is synthesized by hematopoietic cells, megakaryocytes and monocytes.

**Aims:** To understand RUNX1 regulation of *F13A1* expression in platelet/megakaryocyte and the mechanisms and consequences of decreased *F13A1* in RHD.

**Methods:** We performed studies in platelets, HEL cells and human CD34+ cell-derived megakaryocytes including on clot contraction in cells following small inhibitor (si)RNA knockdown (KD) of *RUNX1* or *F13A1*.

**Results:** Platelet *F13A1* mRNA and protein were decreased in our index patient and in two siblings from an unrelated family with RHD. Platelet-driven clot contraction was decreased in the patient and affected daughter. Promoter studies in HEL cells showed that *RUNX1* regulates *F13A1* transcription; RUNX1 overexpression increased and (si)RNA RUNX1 KD reduced *F13A1* promoter activity and protein. Following *RUNX1* or *F13A1* KD clot contraction by HEL cells was decreased as were FXIII-A surface expression, myosin light chain phosphorylation and PAC1 binding upon activation. *F13A1* expression and clot contraction were impaired on *RUNX1* downregulation in human megakaryocytes.

**Conclusions:** RUNX1 regulates platelet-megakaryocyte *F13A1* expression, which is decreased in RHD, reflecting regulation of a coagulation protein by a hematopoietic transcription factor. Platelet and megakaryocyte clot contraction is decreased in RHD, related to multiple impaired mechanisms including *F13A1* expression, myosin phosphorylation and αII_b_β_3_ activation.

**Scientific category** – Platelets and thrombopoiesis

**Essentials:**

- RUNX1 regulates expression of FXIII-A chain (*F13A1)* in megakaryocytes (MK) and platelets.
- Platelet and MK *F13A1* expression and clot contraction are decreased in *RUNX1* deficiency.
- MK clot contraction, myosin phosphorylation and PAC1-binding are impaired in *F13A1* deficiency.
- Defective clot contraction in RHD arises from defects in multiple platelet-MK mechanisms.

## Introduction

RUNX1 is a major transcription factor that regulates hematopoiesis and megakaryocyte and platelet biology [1–4]. Patients with germline *RUNX1* haplodeficiency (RHD) have thrombocytopenia, platelet function defects, and increased risk of myeloid malignancies, referred to as familial platelet disorder with predisposition to myeloid malignancy (FPDMM) [1, 5–8]. The platelet defects in FPDMM include granule deficiencies, impaired platelet aggregation, secretion, protein phosphorylation (myosin light chain, pleckstrin), integrin αII_b_β_3_ activation, and endocytosis [1, 8, 9].

Numerous platelet-MK genes are downregulated [10–12] in FPDMM and we have shown that several, including *MYL9*, *PF4*, *ALOX12*, *PRKCQ*, *ALOX12*, *RAB1B*, *RAB31, PLDN*, and *PCTP,* are direct RUNX1 targets associated with alterations in platelet-MK function [8, 9, 13, 14]. Platelet expression profiling of a FPDMM patient studied by us [10] showed that *F13A1*, which encodes the catalytic A chain (FXIII-A) of plasma and cellular coagulation factor FXIII [15–17], was decreased compared to healthy controls (fold change ratio, 0.30; P=0.006).

FXIII (FXIIIA_2_B_2_) is a transglutaminase composed of a tetrameric complex of homodimers of two A chains with catalytic activity and two B chains (FXIII-B), and induces clot stabilization via cross-linking of fibrin monomers and other proteins [15–18]. Inherited plasma FXIII deficiency causes a major bleeding diathesis with decreased wound healing [16, 17]. FXIII-A (gene *F13A1*) is synthesized in hematopoietic cells, MK and monocytes, and FXIII-B (gene *F13B*) is synthesized by hepatocytes [15–17, 19]. FXIII-A is abundant in platelets (3% by weight) and FXIII-A concentration is ∼100-150-fold higher in platelet cytoplasm than in plasma [16]. Despite being one of the most abundant transcripts in platelets [10, 20] little is known about the regulation of *F13A1* in MK/platelets.

Clot contraction, the volumetric shrinkage of blood clots – plays a major role in hemostasis and thrombosis [18, 21] and is one of the least studied reactions that follow blood clotting and thrombosis [18, 21]. This process is driven by platelet activation mediated alterations in contractile proteins (actin and myosin) that generate mechanical forces, pulling on fibrin attached via the integrin αII_b_β_3_ to the platelet surface to compact the clot. The mechanical properties of fibrin depend on cross-linking by activated FXIII (FXIIIa) [21]. Numerous intracellular proteins involved in clot contraction are cross-linked by FXIIIa [16]. Factor XIII has been implicated in clot contraction [21–24], although some early studies [25, 26] found it to be normal in patients with FXIII deficiency. Clot contraction is impaired in FXIII-A deficient mice [23]. FXIII-A was shown to mediate fibrin translocation to sphingomyelin-rich rafts on platelets to colocalize with myosin and αII_b_β_3_ [22]. Human platelet FXIII-A has been reported to regulate clot contraction [24]; these authors found also that platelet fibrinogen binding, a key event in clot contraction, was decreased in FXIII-A deficient platelets. FXIIIa-crosslinking mediates RBC retention in contracting clots and regulates clot weight [27, 28].

Our present studies provide the first evidence that *F13A1* is transcriptionally regulated by RUNX1 and that FXIII-A is decreased in platelets and MK with RHD. Based on the decreased platelet *F13A1* expression and the platelet abnormalities in agonist-stimulated myosin phosphorylation and αII_b_β_3_ activation described by us [29] in our FPDMM patient, we hypothesized that clot contraction is impaired in *RUNX1* deficiency. Our studies provide evidence for this in platelets, HEL cells and MK with *RUNX1* deficiency and that *F13A1* downregulation is associated with impaired clot contraction. Our studies provide novel information that FXIII-A regulates phosphorylation of myosin light chain (MLC), a key mechanism in clot contraction [18, 21], in megakaryocytic cells. We propose that the impaired clot contraction in *RUNX1* haplodeficiency is due to concurrent defects in multiple mechanisms, including αII_b_β_3_ activation, myosin phosphorylation and FXIII-A expression. Overall, these findings demonstrate regulation of a coagulation factor by a hematopoietic transcription factor, RUNX1.

## Methods

### Reagents and antibodies

Details are provided in Supplemental information.

### Patients with RUNX1 FPDMM

We have previously described [30] the clinical presentation and studies of a male in his 40’s (P1) with a single point mutation in *RUNX1* (c.352-1G>T) in intron 3 at the splice acceptor site for exon 4. We performed studies on this patient and his teenage daughter with the same mutation. We studied two other patients, unrelated siblings (age range 1-10 years) with a germline *RUNX1* mutation (c.508+1G>A). Age- and gender-matched healthy participants were included in this research, approved by the institutional human subject review board, and all participants gave written informed consent.

### Platelets and red blood cells preparation

Whole blood was collected from patients and healthy individuals in one-tenth volume of 3.8% sodium citrate. Platelet-rich plasma (PRP) was prepared by centrifugation at 180g for 20 minutes at room temperature. Platelets were washed twice and pelleted by centrifugation at 600*g* for 20 minutes and resuspended in modified Tyrode’s buffer (pH 7.4); platelet count was adjusted to 3×10^8^/ml. Red blood cells (RBCs) were isolated after removing PRP and buffy coat, RBCs were washed thrice in isotonic solution (0.9% NaCl, 200*g* for 5 minutes), counted with the Hemavet cell counter and used for clot contraction studies in HEL cells or MK at 3–5.0×10^5^ /μl, final concentration [27]. Platelet poor plasma (PPP) was obtained by centrifugation at 3000 rpm for 30 minutes and aliquots stored at -80°C.

### Cell cultures

HEL cells (HEL 92.1.7 from American Type Cell Culture, Rockville, MD) were grown in RPMI-1640 media supplemented with 10% FBS and 100 U/mL Penicillin/streptomycin at 37°C in 5% CO_2_ and megakaryocytic differentiation was induced with PMA (30 nM).

Primary MK were grown *in vitro* from human CD34+ hematopoietic stem cell progenitors (HSPCs) and validated for studies on *RUNX1* deficiency [12, 31]. Briefly, CD34+ cells were infected with shRUNX1 or shNT-lentiviruses expressing a shRNA for either *RUNX1* or a nontargeted sequence as well as mCherry. Cell expressing mCherry were sorted on day 4 and cultured until days 11 to 12 to obtain RUNX-deficient (shRX) and control (shNT) MK used for clot contraction studies.

### Immunostaining

Platelets from the patient P1 and healthy donors were seeded on coverslips coated with human plasma fibronectin and incubated with FXIII-A antibody (Santa Cruz, sc-376312) and imaged by epifluorescence microscopy with a Nikon E100 microscope. Post-acquisition processing and analysis was performed with Adobe Photoshop and NIH-ImageJ and was limited to image cropping and brightness/contrast adjustments applied to all pixels per image simultaneously.

### FXIII plasma levels

Factor XIII (FXIII-A_2_B_2_) was measured in plasma harvested from blood collected into one-tenth volume of 3.8% citrated plasma using a commercially available enzyme-linked immunosorbent assay (Abcam, ab108836).

### Cell extracts and immunoblotting

Lysates from platelets, HEL cells or CD34+-cell-derived MK were lysed in RIPA lysis buffer with protease inhibitor cocktail and subjected to gel electrophoresis on 4-20% Mini-PROTEAN TGX gels, and the protein transferred to PVDF membranes. Membranes were probed with the indicated antibodies listed in Table supplemental (S)1 and IRDye-labeled secondary antibodies and proteins quantified using the Odyssey Infrared Imaging System (Li-Cor, Biosciences, Lincoln, NE).

### Promoter studies

Methods for quantitative real-time PCR, chromatin immunoprecipitation (ChIP) assay, electrophoretic mobility shift assays (EMSA), promoter and plasmid constructs, luciferase reporter assay and studies on the effect of *RUNX1* or *F13A1* downregulation and over-expression are shown in Supplemental Methods.

### Computational Bioinformatics

Potential transcription factor RUNX1 binding sites were predicted by use of TFsearch and TFBIND INPUT (http://www.cbrc.jp/research/db/TFSEARCH.html, http://tfbind.hgc.jp/).

### Clot contraction assays in patients with RUNX1 FPDMM

Clot contraction studies were performed in siliconized glass, aggregometer tubes coated with Sigmacote (Sigma), following the manufacturers recommendation. Whole blood clot contraction (300 µl) was induced by tissue factor (final 1 pM) (American Diagnostics, LLC. Stamford, CT) supplemented with CaCl_2_ (final 10 mM). For clot contraction studies in platelets, 1.5×10^5^ platelets/µl were suspended in healthy donor PPP with added thrombin (5 U/ml) and CaCl_2_ (10 mM final) to induce clotting. Clot contraction tubes were incubated at 37°C in a water bath for up to 120 minutes and images were taken over time. (Apple, iPhone 7). The area of clot sizes relative to initial suspension volumes was determined using ImageJ software (NIH). Clot contraction was calculated as the change in clot area over time relative to initial suspension volume area, and expressed as a percentage of the suspension volume.

### Clot contraction studies in HEL cells and human megakaryocytes

After small inhibitor (si)RNA knockdown of *RUNX1* and *F13A1*, HEL cells were harvested at 48 h post-transfection for clot contraction studies. HEL cells or MK (1.5-2×10^3^/µl) were suspended in Tyrode’s buffer. RBC’s (3–5.0×10^5^/μl) and fibrinogen (0.5 mg/ml) were added, and clotting initiated with 5 U/mL thrombin and CaCl_2_ (10 mM final) at 37°C. Clot contraction was followed for 120 minutes with images obtained at intervals. In studies in HEL cells suspended in PPP no supplemental fibrinogen was added and clotting was initiated with 5 U/mL thrombin and 10 mM CaCl_2_. Where indicated, HEL cells were pre-incubated for 20 minutes at 37°C with an inhibitor of myosin II activity (blebbistatin 200 μM) [21], αII_b_β_3_ (eptifibatide acetate 125 μM) or FXIIIa activity (T101 100 μM) [27] prior to clot contraction studies .

### FXIII-A expression on platelets and HEL cells by flow cytometry

To evaluate cellular FXIII-A, platelets were exposed to agonists (ADP 20 μM, collagen 2.5 µg/mL, SFLLRN 10 μM, A23187 10 μM alone or in combination) for 8 minutes and incubated with AlexaFluor-647®-conjugated anti-FXIII-A antibody (Abcam, ab225018) at RT for 45 minutes without stirring. HEL cells (1×10^6^/ml) were activated with a combination of convulxin (100 ng/ml) and thrombin (5 U/ml) for 20 minutes prior incubation with FXIII-A antibody. The reaction was stopped with 2% paraformaldehyde, cells washed 3 times with PBS and analysed on a Becton Dickinson, BD^TM^ LSRII flow cytometry within four hours. The cytometer was calibrated before the assay using BD CS&T research beads.

### Studies on MLC phosphorylation in HEL cells

PMA-HEL cells transfected with siRNA KD of *F13A1* or *RUNX1* were treated with 5 U/ml thrombin and 25 mM CaCl_2_ for 10 minutes at 37°C. Cell lysates were subjected to immunoblotting using antibodies against MLC, phosphorylated MLC (pMLC) and actin.

### Statistical methods

Results are expressed as mean ± SEM. Differences were compared using Student’s t-test or one-and two-way ANOVA, using GraphPad Prism, version 8 (GraphPad Software) and considered significant at P< 0.05.

## Results

### Platelet *F13A1* expression is decreased in FPDMM patients

Prior expression profiling studies [10] using Affymetrix U133 Gene Chips showed that platelet *F13A1* expression was decreased in our FPDMM patient compared with healthy controls (fold change = 0.30; P=0.006). We validated this finding with real-time qPCR; platelet *F13A1* mRNA expression was lower than in healthy, age and gender-matched subjects (Figure 1A). It was also lower compared to healthy controls in 2 siblings with FPDMM who were unrelated to our index patient P1 (Figure 1A). Decreased platelet *F13A1* expression has also been observed by others in 2 FPDMM patients with different *RUNX1* mutation than in our patients. [11].

**Figure 1.**
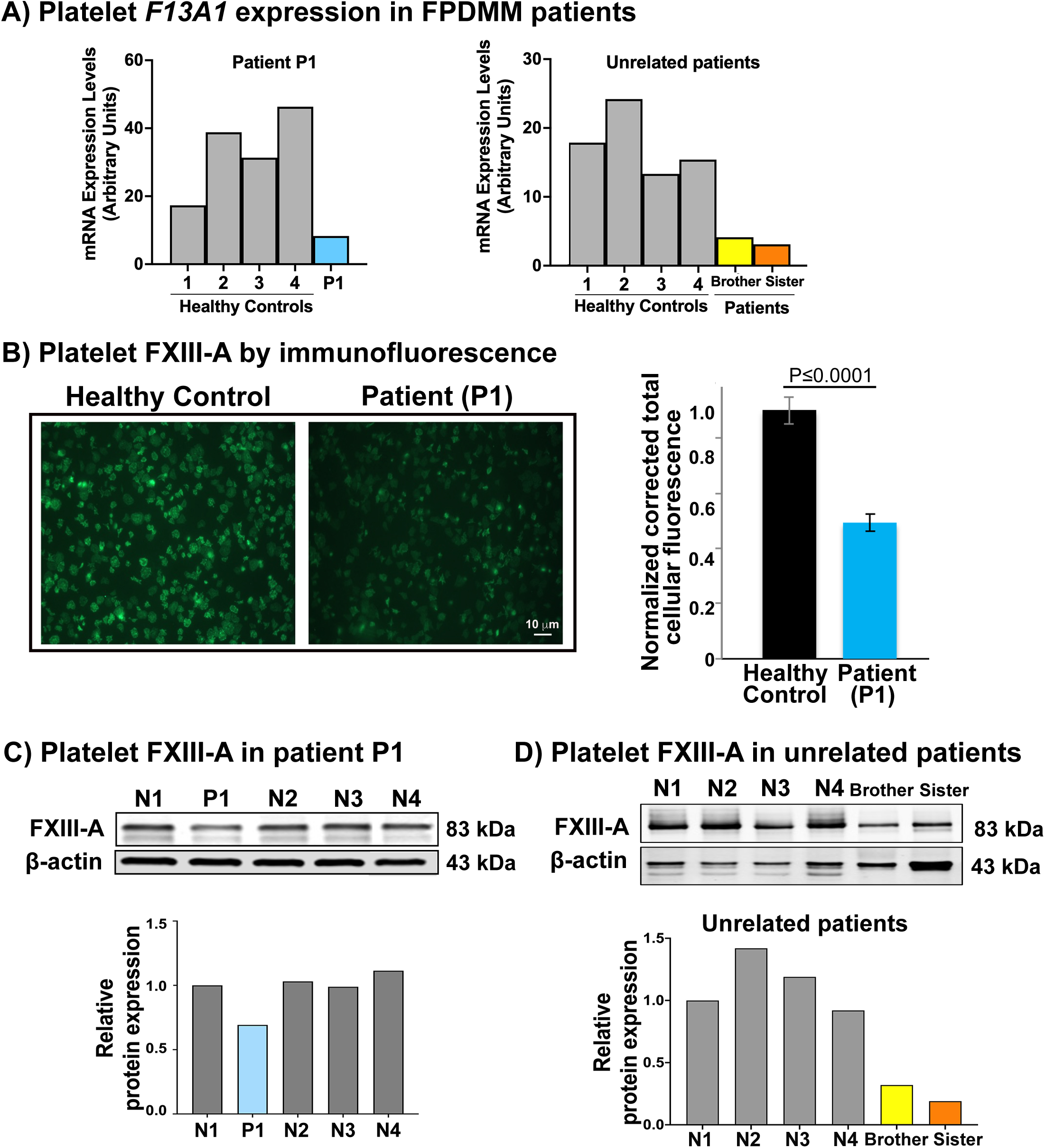
*F13A1* expression in platelets and plasma in FPDMM patients. **A)** Platelet *F13A1* mRNA levels by qPCR in healthy controls and patient P1 (father) and 2 unrelated patients (brother and sister). Shown are mRNA levels normalized to GAPDH. **B)** FXIII-A by immunostaining in platelets from the patient P1 and healthy control. Bar graph shows normalized corrected total cellular fluorescence (mean ± SEM) calculated from images. **C)** Immunoblot analysis showing FXIII-A expression in platelet lysates from patient P1 and 4 healthy controls with β-actin as loading control. The bar graphs below show FXIII-A quantification normalized to actin. **D)** Immunoblot analysis showing FXIII-A expression in platelet lysates from 2 unrelated siblings and 4 healthy controls. The bar graphs below show FXIII-A quantification normalized to actin.

We evaluated platelet FXIII-A expression in of our index patient by immunostaining. The corrected total cellular fluorescence (CTCF) was decreased ∼ 50% in the patient’s platelets compared to age and gender-matched healthy control (Figure 1B), indicating decreased FXIII-A protein levels in platelets from the patient. Immunoblotting of platelet lysates confirmed decreased FXIII-A levels in the patient (on two different visits) (Figure 1C) and in two unrelated siblings as compared to healthy controls (Figure 1D). Plasma FXIII measured using an ELISA in the index case and his daughter with FPDMM were within the range observed in 17 healthy donors (not shown).

### RUNX1 regulates *F13A1* in megakaryocytic HEL cells

*In silico* analyses of the 5’ upstream region of *F13A1* showed 7 RUNX1 consensus sites within 545 bp from the ATG start codon (Figure 2A). ChIP studies using PMA-treated HEL cells and RUNX1 antibody indicated that RUNX1 binds in the region encompassing site 4 and sites 5, 6, 7 (Figure 2B). EMSA studies showed specific binding of nuclear protein to labeled DNA probes with sites 1, 4, and combined 5, 6 and 7. RUNX1 antibody produced a “supershift” of the corresponding bands or abolished the band indicating RUNX1 as the protein binding the DNA (Figure 2C). In luciferase reporter studies mutagenesis of RUNX1 binding sites 3, 4, 6, and 7 decreased while mutation of site 5 increased *F13A1* promoter activity, indicating that these binding sites are functional (Figure 2D). Overexpression of *RUNX1* (isoform B) [20] increased *F13A1* promoter activity and FXIII-A protein levels (Figure 2E) while siRNA RUNX1 knockdown reduced *F13A1* promoter activity and FXIII-A protein (Figure 2F). Together, these studies indicate that RUNX1 regulates *F13A1* transcription.

**Figure 2.**
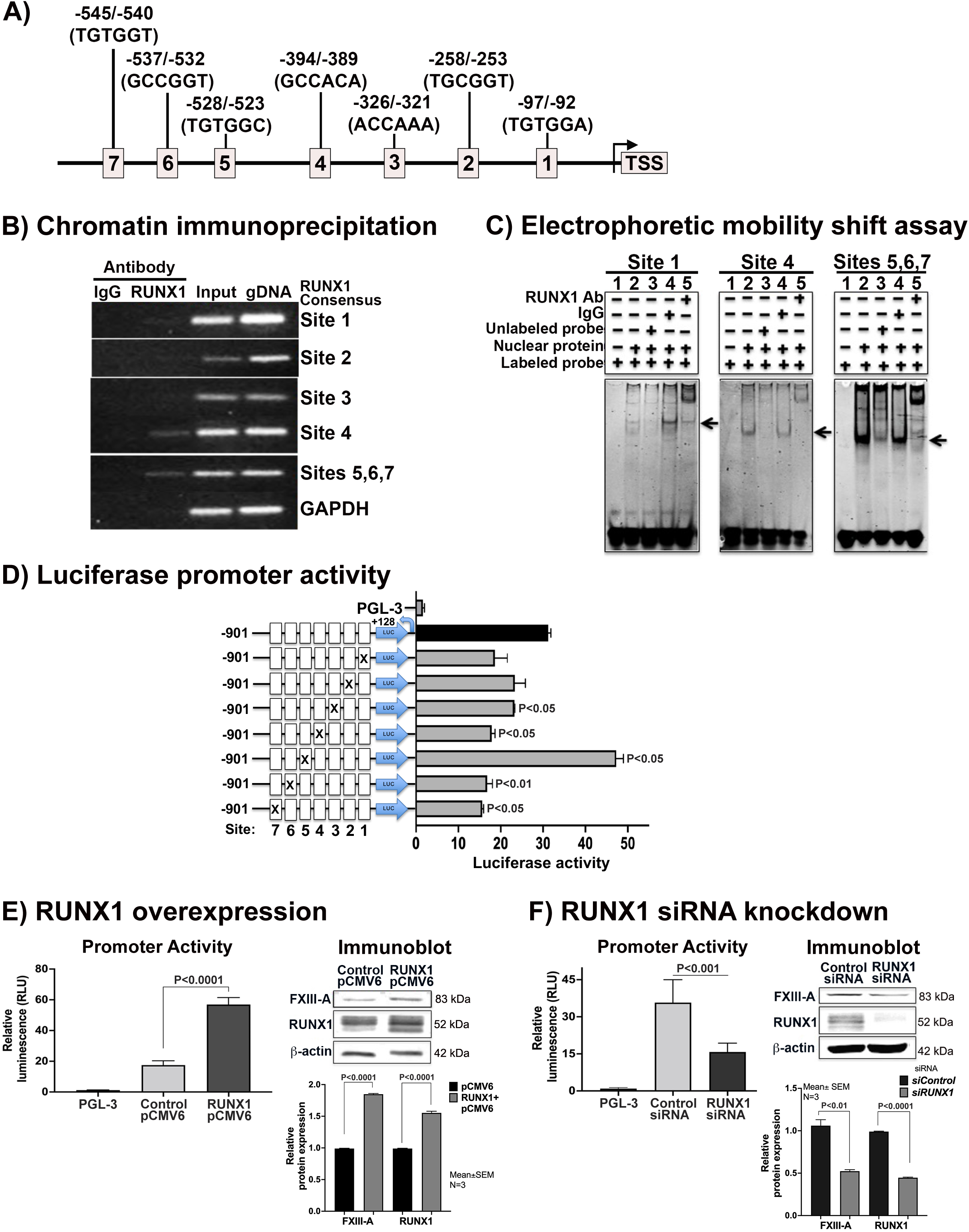
Binding of RUNX1 to *F13A1* promoter region and functional activity in PMA-treated HEL cells. **A)** The 5’ upstream region of *F13A1* with seven RUNX1 consensus-binding sites. **B**) Binding of RUNX1 to *F13A1* promoter region by ChIP using control IgG or RUNX1 antibody. PCR was performed with primers designed to amplify *F13A1* promoter region encompassing RUNX1 binding sites 1 through 7 and GAPDH. Shown are PCR amplification of control IgG and RUNX1 (column 1 and 2), total input DNA and genomic DNA (column 3 and 4) respectively. **C**) Binding of RUNX1 to *F13A1* promoter region by EMSA. Shown is EMSA using oligonucleotide probe with RUNX1 binding site 1, 4, and probe with sites 5, 6, and 7, and nuclear extracts from HEL cells. For each panel, lane 1 is probe alone; lane 2, probe with nuclear extract; lane 3, competition with ×100 molar excess unlabeled probe; lane 4, effect of control IgG; and lane 5, effect of RUNX1 antibodies. Arrows indicate areas where the band was supershifted or competed by the RUNX1 antibodies. The nucleotide sequences of the probes are shown in Table S5 in the supplement. **D**) Luciferase reporter studies on the *F13A1* promoter. Shown are luciferase promoter activity at 24 h of the wild-type construct and effect of mutating each of the 7 RUNX1 consensus binding sites in the 5’ upstream region of *F13A1*. The ratios of luciferase to renilla activity of the wild-type construct and constructs with each RUNX1 consensus site mutated are shown. Site-directed mutagenesis of the RUNX1 binding sites on *F13A1*promoter decreased promoter activity with the exception of site 5, which seemed to exhibit negative regulation. **E)** Effect of *RUNX1* overexpression on *F13A1* promoter activity. PMA-treated HEL cells were transfected with the empty pCMV6 vector or RUNX1B-pCMV6 expression vector together with *F13A1*-promoter-PGL3-luciferase vector. Shown are luciferase promoter activities at 24 h compared to that of cells transfected with promoterless PGL3-basic vector (mean ± SEM of 3 experiments) and immunoblots of cell lysates for FXIII-A and RUNX1 with β-actin as loading control with quantification bar graphs at the bottom. **F)** Effect of RUNX1 siRNA on *F13A1* promoter activity. PMA-treated HEL cells were cotransfected with control or RUNX1 siRNA together with *F13A*1-promoter-PGL3 luciferase reporter constructs. Reporter activity was measured at 24 h. The mean ± SEM of 3 experiments performed in triplicate are shown. Also shown are immunoblots of cell lysates for FXIII-A and RUNX1 with β-actin as loading control; bar graphs show quantification from 3 experiments.

### Studies on clot contraction studies in whole blood and platelets suspended in PPP

In 2 FPDMM patients (father P1 and daughter P2) we assessed clot contraction over 120 minutes in whole blood and by washed platelets suspended in PPP (Figure 3). In whole blood clotting was initiated with tissue factor (1 pM) and CaCl_2_ (10 mM). In both patients clot contraction appeared similar to that in healthy controls (Figure 3 B and D top rows). In parallel studies in washed platelets suspended in PPP clotting was initiated with 5 U/ml thrombin in the presence of CaCl_2_. Platelet clot contraction in both patients was markedly delayed over 60-120 minutes, with smaller clots and visually less dense clots compared to those in the healthy controls (Figure 3 B and D, bottom rows).

**Figure 3.**
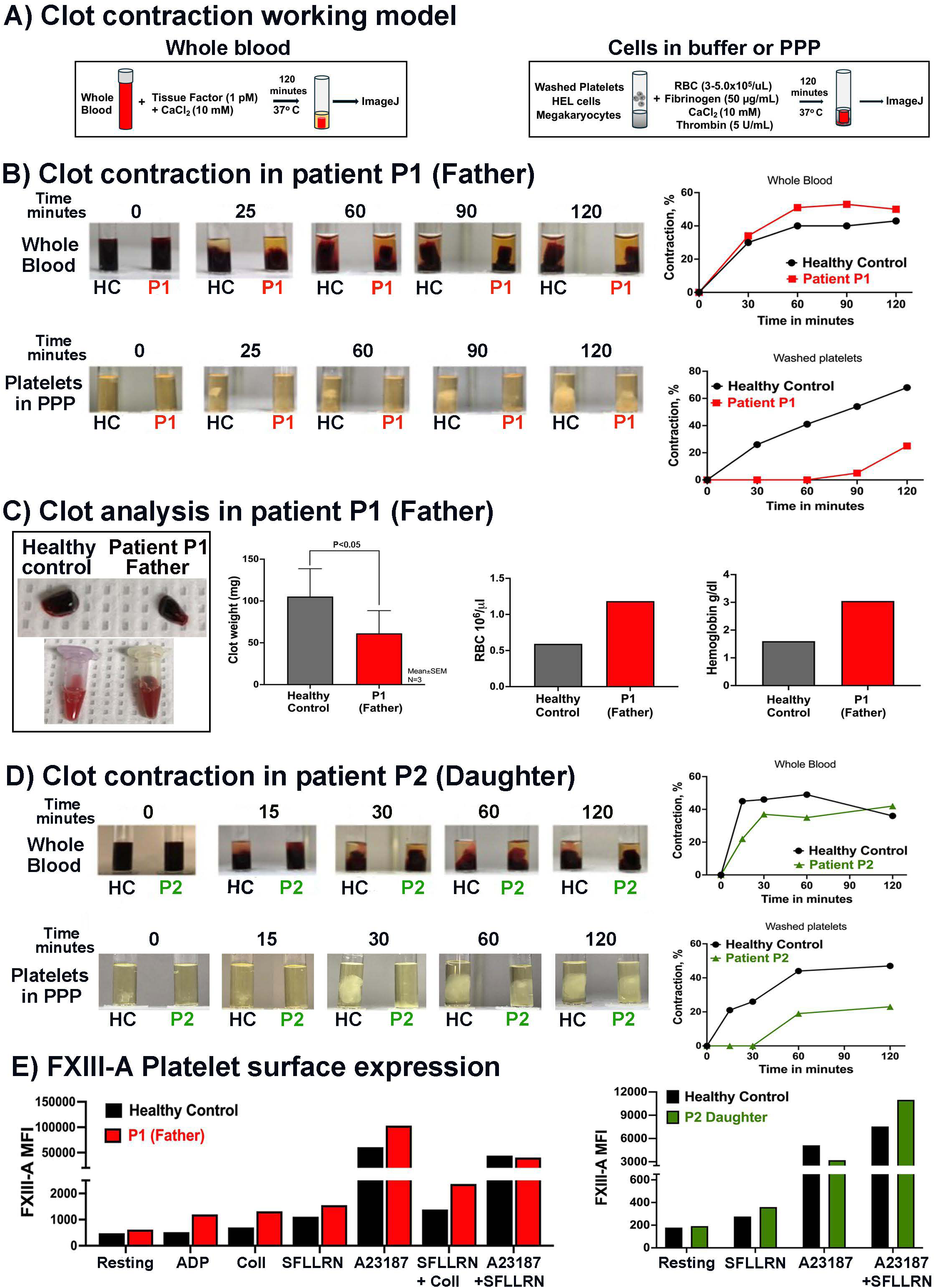
Clot contraction is impaired in FPDMM patients (father and daughter) as compared to controls. **A)** Clot contraction working model for studies in whole blood and using cell suspensions in buffer. **B)** Clot contraction studies in a healthy donor and patient P1 (father) in whole blood (top row of tubes) and in washed platelets (3×10^8^/ml) suspended in healthy donor platelet poor plasma (PPP) (lower row) over 120 minutes. Graphs on the right of each row of tubes show percent clot contraction over time, **C)** Shown are pictures of clots from healthy donor and patient P1 (father) at 120 minutes and the serum after removal of the clots; representative of two independent experiments. Bar graphs show clot weights from 3 independent experiments, and RBCs counted and hemoglobin concentration in the serum from two separate visits. **D)** Clot contraction studies performed in healthy control and patient P2 (daughter) in whole blood (top row of tubes) and using washed platelet suspended in PPP (bottom row). The graphs next to the tubes show the percent contraction over time. **E)** FXIII-A surface expression in resting and agonist stimulated platelets from healthy volunteers (black bars) and patients (P1: red and P2: green) with FDPMM by flow cytometry. Data are shown as MFI (mean fluorescent intensity).

In studies done in whole blood we evaluated the clot weights. Clots from the father weighed less than clots from control subjects (Figure 3C). FXIII regulates retention of RBC in blood clots and it is decreased in FXIII deficiency [27]. On two separate visits, we measured the RBCs and the hemoglobin concentration in the serum after removal of the clots. In the father, the number of RBCs and hemoglobin concentration in the serum were higher compared to that in the healthy control, suggesting impaired RBC retention (Figure 3C). In the daughter, the clot weight and RBCs in the serum were no different from the control (not shown).

### Expression of FXIII-A on platelet surface on activation by flow cytometry

Resting platelets have negligible FXIII-A on their surfaces and it is enhanced upon activation with strong agonists [32, 33]. FXIII-A surface expression in resting platelets was very low in healthy subjects and patients. It increased upon activation with A23187 alone or in combination with SFLLRN but not with ADP, SFLLRN or collagen alone (Figure 3E). In both the father and the daughter, platelet FXIII-A surface expression was similar to that in the controls in the resting state and following activation (Figure 3E). Only a small fraction of the total FXIII-A is expressed on platelet surface upon activation [32, 34]; the bulk is retained in the cell [34]. Thus, the partial FXIII-A deficiency in RHD may not alter surface expression.

### Clot contraction is impaired in *RUNX1-* and *F13A1-* deficient HEL cells

PMA-treatment induces megakaryocytic transformation of HEL cells with increased expression of αII_b_β_3_ and FXIII-A [20] These cells express GPIb, produce thromboxane A2 and demonstrate fibrinogen-binding and myosin phosphorylation, two major players in clot contraction. We assessed clot contraction in HEL cells with and without PMA treatment (Figure 4A). RBCs are incorporated and retained in the contracting clots by the action of FXIII-A [27]; therefore, we added RBCs to the cell suspensions. Compared to untreated cells clot contraction was increased by PMA-treated HEL cells, (Figure 4A), which reflects the effect of megakaryocytic transformation. In general, clot contraction was relatively less intense in control PMA-treated HEL cells compared to that in PRP or WB (not shown).

**Figure 4.**
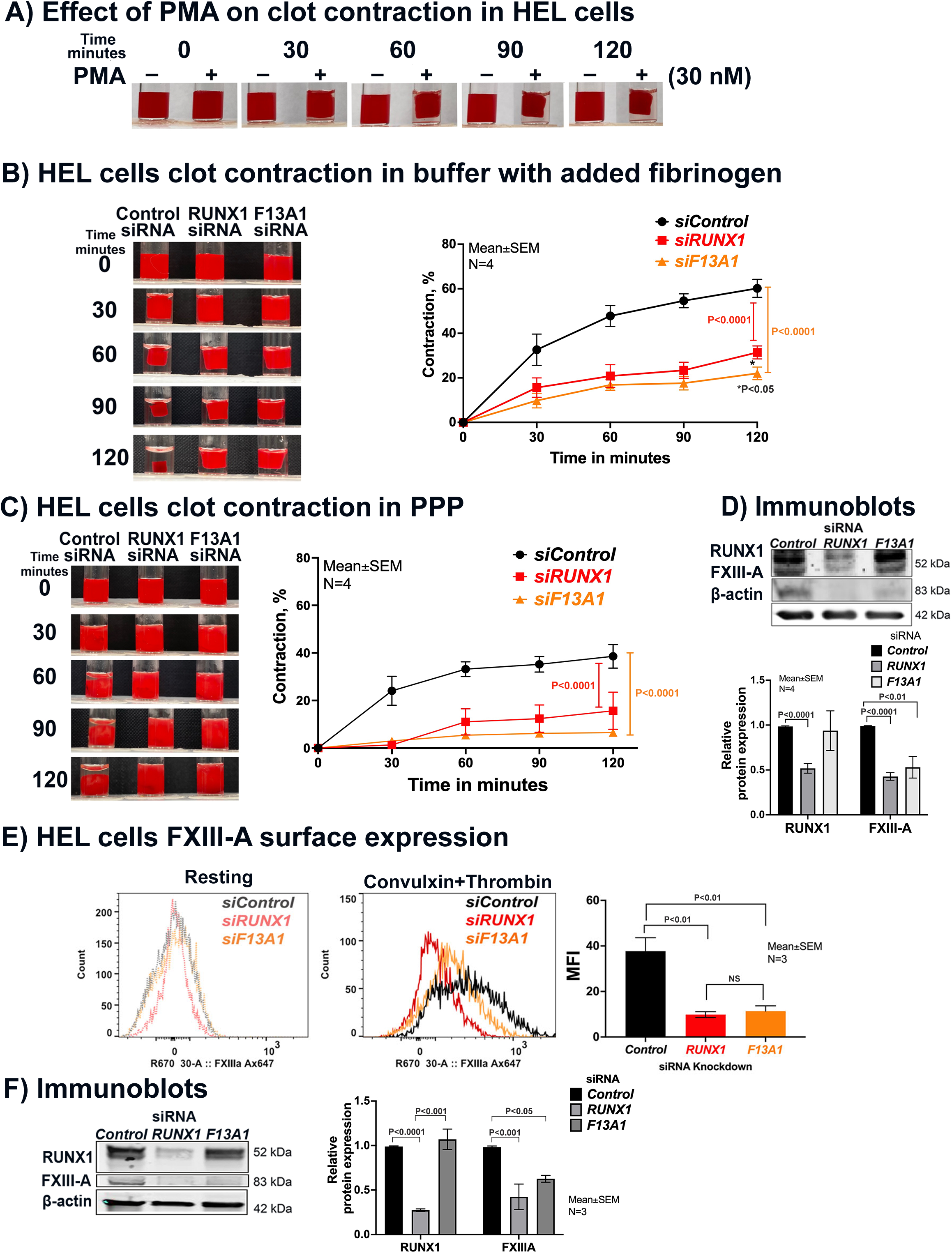
Clot contraction in megakaryocytic HEL cells and CD34+ megakaryocytes. **A**) Clot contraction in HEL cells without (-) or with (+) PMA treatment. HEL cells grown with 30 nM PMA for 4 days were used to evaluate clot contraction for up to 120 minutes. Shown representative of 2 separate experiments. **B)** Clot contraction in PMA-treated HEL cells following siRNA KD of *RUNX1* or *F13A1.* Cells were suspended in buffer with added RBCs and 50 μg/ml fibrinogen and clotting initiated with 5 U/ml thrombin and 10 mM CaCl_2_. Line graph represents percent contraction up to 120 minutes; shown are mean ± SEM of 4 independent experiments. The extent of contraction was determined from the pictures using ImageJ software. **C)** Clot contraction in PMA-treated HEL cells suspended in PPP with added RBCs and clotting initiated with 5 U/ml thrombin and 10 mM CaCl_2_. Shown is a representative experiment. Line graph shows mean ± SEM of extent of clot contraction over 120 minutes from 4 independent experiments. **D**) Immunoblots of HEL cell lysates for RUNX1 and FXIII-A in control cells and following siRNA KD of *RUNX1* or *F13A1* with β-actin as loading control. The bar graphs show quantification of protein bands normalized to β-actin (n=4). **E)** FXIII-A surface expression in resting and convulxin + thrombin activated HEL cells on *RUNX1* and *F13A1* siRNA knockdown by flow cytometry. Bar graph shows MFI from three independent experiments (mean ± SEM). **F)** Shown are immunoblots of cell lysates for RUNX1, FXIII-A and β-actin as a loading control and quantification of protein bands normalized to β-actin (n=3).

Clot contraction was evaluated in PMA-treated HEL cells following siRNA KD of *RUNX1 or F13A1*. Cells were suspended in buffer with added fibrinogen (50 μg/ml) or in PPP and clotted in the presence of added RBCs with thrombin and 10 mM CaCl_2_. Compared to control cells, the clot contraction was reduced on *RUNX1* or *F13A1* knockdown; this was noted in cells suspended in either buffer or in plasma (Figure 4B, C). Together, they suggest that cellular FXIII-A contributes to clot contraction and this is noted even in the presence of plasma FXIII. Of note, in these studies the KD of *RUNX1* or *F13A*1 was only partial (∼50-60%) to mimic what is noted in patients with RHD (FPDMM) who have a heterozygous abnormality [1, 5, 8] (Figure 4D, F).

### FXIII-A surface expression on HEL cells upon activation by flow cytometry

Resting platelets have minimal FXIII on the platelet surface and activation leads to enhanced surface expression [32, 33]. Resting HEL cells had negligible surface FXIII-A and activation with convulxin (100 ng/ml) plus thrombin (5 U/ml) resulted in an increase in FXIII-A surface expression (Figure 4E). siRNA KD of *RUNX1* or *F13A1*, markedly decreased FXIII-A surface expression in activated HEL cells compared to control cells (Figure 4E).

### Effect of *RUNX1* KD in MK differentiated from human CD34+ HSPCs

To corroborate the findings in HEL cells (Figure 4), we performed studies in human MK derived from HSPCs [12, 31]. By scRNA-sequencing, cells with *RUNX1* KD (shRX) had reduced expression of *F13A1* compared to control cells (shNT) (Figure 5A). Compared to control MK (shNT), clot contraction was reduced on *RUNX1* knockdown (shRx) (Figure 5B) in cells suspended in buffer with added fibrinogen and RBCs and clotting initiated with thrombin (5 U/ml). These recapitulate the findings in HEL cells.

**Figure 5.**
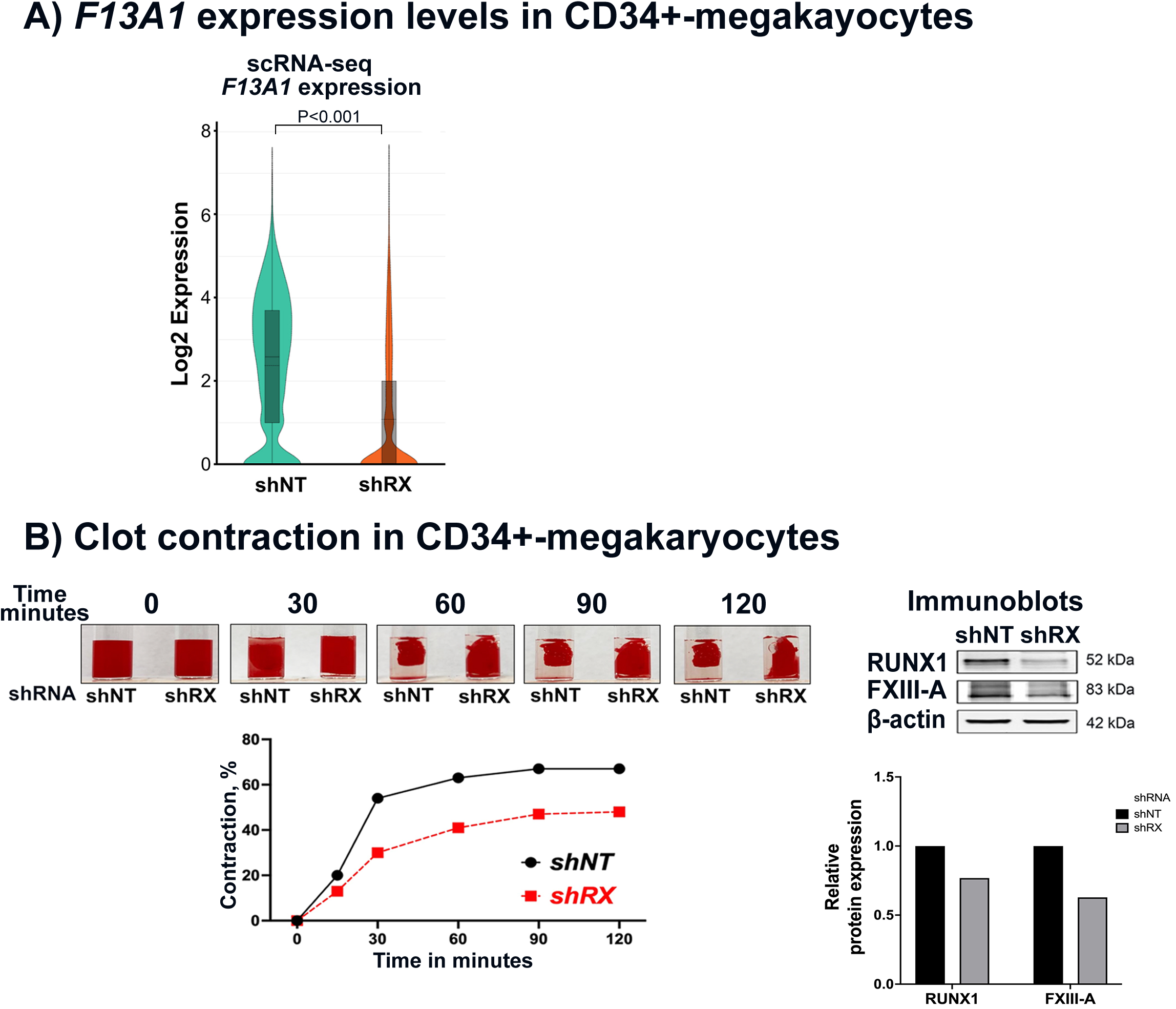
*F13A1* expression and clot contraction in short hairpin RNA *RUNX1* KD (shRX) in CD34+-derived megakaryocytes. A) Violin plot with box plots illustrating the distribution of log2 expression levels of *F13A1* gene in CD34+-MK cells differentiated for 14 days, comparing single cells from shRNA-NT (shNT) and shRNA-RUNX1 (shRX) groups, based on single-cell RNA-seq data [12, 31]. The central line within each box represents the median expression value, with the lower and upper edges of the box indicating the first (Q1) and third (Q3) quartiles, respectively. The whiskers extend to the minimum and maximum observed values. **B)** Shown is clot contraction up to 120 minutes in shRX and control shNT megakaryocytes in buffer with added RBCs, fibrinogen, and CaCl_2_; clotting was initiated with thrombin Pictures are representative of two independent experiments. Line graph below shows clot contraction over time up to 120 minutes. Immunoblots show RUNX1 and FXIII-A protein expression and β-actin as loading control. Bar graph shows protein band intensities normalized to β-actin.

### Agonist-stimulated PAC1-binding and MLC phosphorylation are decreased in *RUNX1-* and *F13A1-*deficient HEL cells

Two major players in clot contraction are the surface αII_b_β_3_ complexes that function as fibrin(ogen) binding sites upon cell activation and the phosphorylation of myosin light chain [18, 21]. *RUNX1* or *F13A1* KD did not alter αII_b_β_3_ complexes on HEL cells when assessed using an antibody (CD41-FITC) that binds the integrin in resting cells (Figure 6A). Upon thrombin activation, whereas PAC1 binding indicating αII_b_β_3_ activation was strongly induced in control cells, it was decreased following *RUNX1* KD, in line with our studies in HEL cells [9] and FPDMM platelets [29]. Unexpectedly, PAC1-binding was decreased on *F13A1* KD compared to control cells (Figure 6B). We have previously shown that thrombin stimulated MLC phosphorylation is decreased in FPDMM platelets [29] and in HEL cells on *RUNX1* KD [20], secondary to decreased MLC protein level because RUNX1 regulates *MYL9* [35]. We now found thrombin stimulated MLC phosphorylation was decreased on *F13A1* KD (Figure 6C), suggesting that FXIII-A regulates MLC phosphorylation. In striking contrast to decreased MLC levels observed with *RUNX1* KD, total levels of MLC by immunoblotting were normal on *F13A1* KD. These results suggest that the decreased MLC phosphorylation observed with *RUNX1* KD and *F13A1* KD arise by different mechanisms.

**Figure 6.**
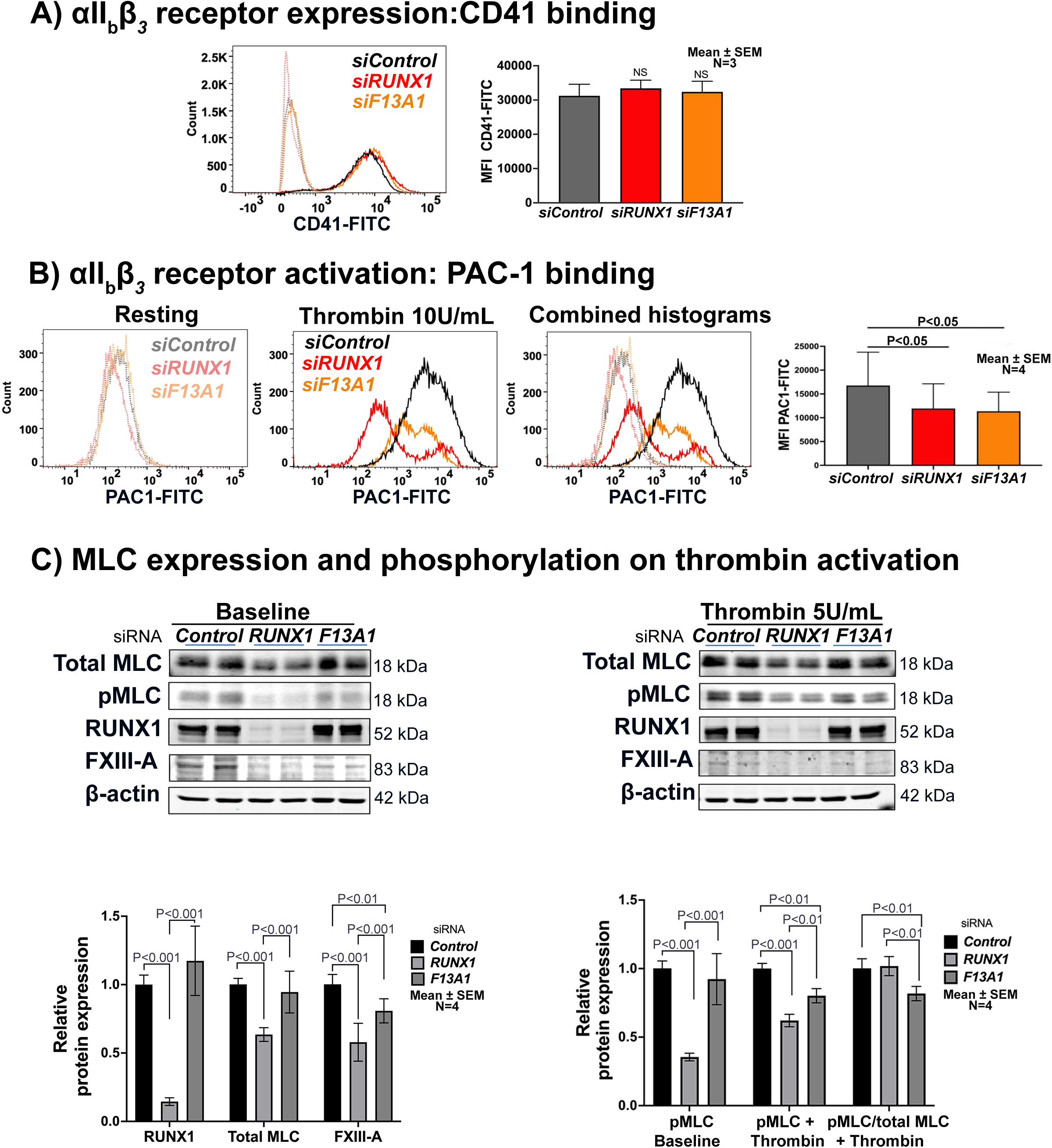
Effects of *RUNX1* and *F13A1* siRNA kD on αII_b_β_3_ expression and activation and MLC expression and phosphorylation in megakaryocytic HEL cells. **A)** Histograms show αII_b_β_3_ receptor expression on *RUNX1* (red)- and *F13A1* (orange)-deficient HEL cells as compared to control cells (black) by flow cytometry with CD41-FITC antibody. Bar graph (right) shows CD41 expression (MFI), representative of three independent experiments (mean ± SEM). The differences between the 3 groups were not statistically significant. **B)** Effect of siRNA kD of *RUNX1* (red) or *F13A1* (orange) on αII_b_β_3_ receptor upon activation (PAC1 binding) with thrombin (10 U/mL) in HEL cells as compared to control cells (black). Shown are representative histograms with PAC1-FITC antibody on resting and thrombin activated cells and combined histograms. Bar graph shows MFI of four independent experiments (mean ± SEM). **C)**. MLC expression and its phosphorylation upon thrombin activation in HEL cells in the baseline resting state (left panel) and following thrombin activation (right panel). Left panel: Representative immunoblots of total MLC, phospho-MLC (pMLC), RUNX1, FXIII-A and β-actin protein expression in control cells and *RUNX1-* and *F13A1-* deficient cells in the resting state (baseline). Right panel: shows immunoblots after thrombin activation (5 U/mL). Bar graphs under the left immunoblot show relative protein expression of RUNX1, total MLC and FXIII-A at baseline. The bar graphs on the right show pMLC at baseline (unactivated) and after thrombin activation in control cells and in cells with KD of *RUNX1* or *F13A1.* The last set of bars show pMLC expressed as a fraction of the total MLC present in control cells and cells with *RUNX1* or *F13A1* KD. Data are shown as mean ± SEM (N=4).

### Effect of inhibitors on clot contraction in HEL cells

To corroborate the findings in HEL cells, we studied the effect of inhibitors of αII_b_β_3_ (eptifibatide) [36], myosin light chain (blebbistatin) [36] or FXIIIa (T101) [23, 28] and each inhibited clot contraction by HEL cells (Figure 7A). These results demonstrate that HEL cell clot contraction is dependent on the respective mechanisms inhibited.

**Figure 7.**
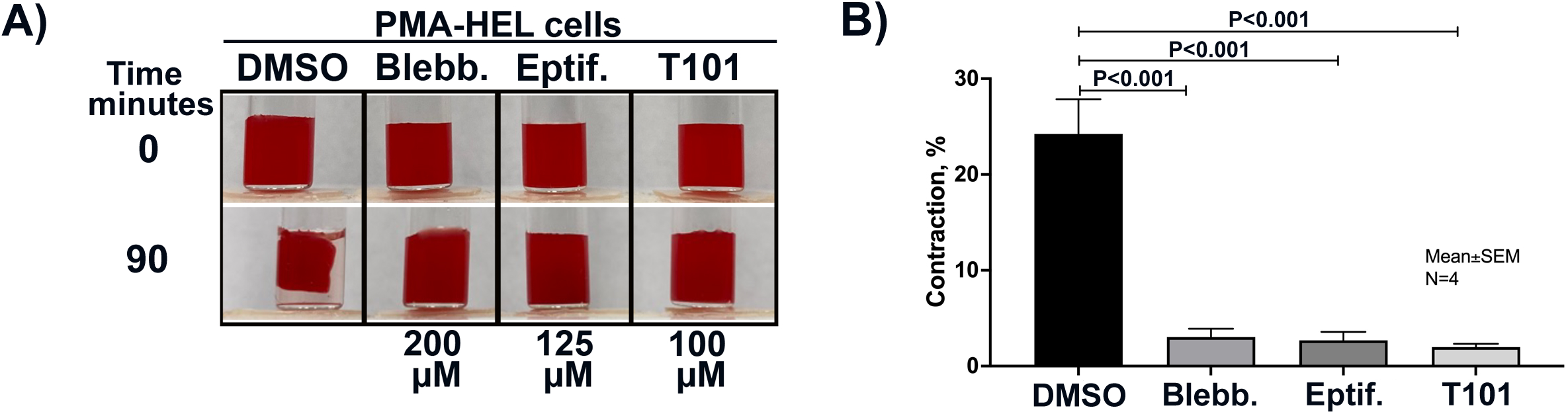
Inhibition of αII_b_β_3_, myosin light chain or FXIIIa inhibits clot contraction in HEL cells. PMA treated-HEL cells suspended in buffer were incubated for 20 minutes with inhibitors of αII_b_β_3_ (Eptif = eptifibatide), MLC (Blebb= blebbistatin), or FXIIIa (T101) and clot contraction was initiated with thrombin (5 U/ml) and CaCl_2_. Bar graphs show mean ± SEM of percent contraction at 90 minutes evaluated by ImageJ software (n=4).

## Discussion

Our studies provide the first evidence that *F13A1* expression is decreased in platelets of FPDMM patients studied by us (Figure 1A-D), and in *RUNX1*-deficient HEL cells (Figure 4D-F) and MK (Figure 5A), and that *F13A1* is a direct transcriptional target of RUNX1 in platelets/MK. Decreased platelet *F13A1* mRNA was found in 2 FPDMM patients studied by others [11] indicating that our findings may apply to FPDMM patients beyond those studied by us. Moreover, we previously found [20] that platelet *F13A1* transcript levels in healthy donors, measured by RNA-seq, correlate with those of total *RUNX1* and *RUNX1B* isoforms. This provides additional and *in vivo* support to the conclusion that RUNX1 regulates *F13A1.* FPDMM patients have a heterozygous *RUNX1* mutation and platelet *F13A1* is decreased but not absent (Figure 1A, B).

FXIII induces clot stabilization via cross-linking of fibrin monomers and other proteins [16, 21, 22, 24]. Our studies provide evidence that platelet-driven clot contraction is impaired in FPDMM patients studied (Figure 3B-D). These findings are corroborated by the findings of impaired clot contraction in *RUNX1*-deficient HEL cells and human MK, and in *F13A1-*deficient HEL cells. FXIII regulates retention of RBCs in the clots [27] and the clot weights are proportional to RBCs retained [27]. In the father, the clot weights were lower than in the controls and the non-retained RBCs in the serum were higher, suggesting an abnormality in RBC retention (Figure 3C) and reflecting on another FXIII function.

In initial studies clot contraction was increased in PMA-treated cells compared to untreated cells (Figure 4A), indicating the effect of megakaryocytic differentiation, which is associated with increased expression of αII_b_β_3_ and other processes. In our previous studies *F13A1* expression in HEL cells also increased with PMA-treatment [20]. In general, the extent of clot contraction was less with HEL cells and MK than in platelets (not shown). Our studies show that both clot contraction and *F13A1* expression are decreased in *RUNX1* deficient HEL cells (Figure 4B-C) and human MK (Figure 5B).

Contraction was impaired in *F13A1*-deficient HEL cells suspended in buffer or plasma (Figure 4B-C). It was also decreased by a transglutaminase inhibitor (Figure 7). The impaired clot contraction in *F13A1-*deficient HEL cells suspended in buffer supports the conclusion that cellular FXIII (in distinction from plasma FXIII) plays a role in clot contraction.

Clot contraction is driven by multiple mechanisms: the intracellular contractile forces generated by the actin-myosin machinery, MLC phosphorylation, fibrin(ogen) binding to activated αII_b_β_3_ complexes on platelet surface, FXIIIa mediated cross-linking of fibrin (and intracellular proteins), and translocation of fibrin-myosin-αII_b_β_3_ complex to the phosphatidylserine (PS)-rich lipid rafts in activated cells [18, 21, 22, 24]. What are the mechanisms leading to the impaired clot contraction in platelets and MK with *RUNX1* deficiency, where numerous genes and processes are downregulated [8, 10, 11]? We posit that the impaired clot contraction is related to defects in multiple cellular mechanisms that we have shown in *RUNX1* deficiency, including decreased expression of RUNX1-target *MYL9* (MLC) and its phosphorylation [9, 20, 29, 35] and αII_b_β_3_ activation (PAC-1 binding) [29], and now the decreased FXIII-A. Our studies with inhibitors support the role of these mechanisms in clot contraction; inhibitors of αII_b_β_3_ (eptifibatide), myosin light chain (blebbistatin) and FXIII (T101) all individually inhibited clot contraction in HEL cells (Figure 7A).

One of the earliest abnormalities demonstrated by us in FPDMM platelets is impaired thrombin-induced MLC phosphorylation [29] due to decreased expression of *MYL9* [10]. We now show this occurs in *RUNX1* deficient HEL cells as well (Figure 6C). Both FPDMM platelets [29] and RUNX-deficient HEL cells (Figure 6A) have impaired αII_b_β_3_ activation (Figure 6B), indicating impaired fibrin(ogen) binding to these cells. Thus, in both cells 2 major mechanisms that regulate contraction are impaired. Finally, FXIIIa crosslinks fibrin and regulates translocation of fibrin-myosin-αII_b_β_3_ complex to the PS-rich lipid rafts to contribute to clot contraction [22]. We show that FXIII-A is decreased in RUNX1-deficient platelets, HEL cell and MK (Figures 1, 4D and 5A) and that *F13A1* KD in HEL cells impairs clot contraction (Figure 4 B, C). Overall, multiple abnormalities contribute to the abnormal clot contraction in RHD.

Our studies provide novel information on FXIII-A functions in MK. These include MLC phosphorylation and PAC1 binding on activation, which were both decreased on *F13A1* KD in HEL cells (Figure 6C). The latter implicates FXIII-A in modulating αII_b_β_3_ function and is in line with platelet studies showing that FXIIIa inhibitors blunt fibrinogen binding and aggregation [24]. Our studies suggest that the decreased MLC phosphorylation following *RUNX1* and *F13A1* KD arise from distinct mechanisms. With *RUNX1* KD the impaired MLC phosphorylation (Figure 6C) reflects the decreased MLC protein, because *MYL9* is a direct RUNX1 target [9, 35]. In contrast, with *F13A1* KD, MLC protein levels are unaffected although its phosphorylation was decreased (Figure 6C). These findings suggest that other mechanisms (possibly involving the kinases) are responsible for the impaired MLC phosphorylation with *F13A1* KD.

The cellular source of plasma FXIII-A subunit is accepted to be hematopoietic [15–17] although the contribution of specific cells in humans remains unclear and MK as a major source has been questioned in studies in mouse models [19]. Platelet FXIII-A constitutes a large part of circulating FXIII [16], and we show that *F13A1* expression is regulated in MK by RUNX1 with decreased platelet levels in FPDMM. Studies are needed in a large cohort FPDMM patients to clarify whether altered *RUNX1* regulation of *F13A1* in MK-platelet has an impact on plasma levels. These are outside the scope of the present project.

One potential limitation of our studies is the number of FPDMM patients studied, which precludes extrapolating them to the overall FPDMM patients. Decreased platelet *F13A1* mRNA has been observed in 2 other patients by Palma et al [11]. Studies in a large cohort of FPDMM patients are needed to establish the prevalence of impaired clot contraction. In FPDMM, multiple platelet-MK responses and processes are recognized to be altered [1, 8–11]. Our studies extend these to encompass impaired clot contraction.

The molecular mechanisms of clot contraction in platelets and MK are similar or identical [37]. Studies of Kim et al [37] using MK derived from induced pluripotent stem cells (IPSC) and our studies advance MK as a model to study platelet clot contraction. These cells can be genetically modified to study genetic disorders and gene variants, as done by us. MK clot contraction may be physiologically relevant. MKs are present in the lung vasculature and lead to substantial intravascular platelet production [38, 39]. Activated intravascular MK may promote contraction of blood clots to reinforce a hemostatic plug or remodel thrombosis ([37] and references therein).

Our findings on RUNX1-regulation of *F13A1* may be relevant beyond impaired hemostasis in FPDMM, to cardiovascular disease. We have recently shown [20] that *F13A1* transcripts in whole blood are associated with an increased incidence of death or myocardial infarction in patients with cardiovascular disease (CVD) [20]; our previous studies have associated *RUNX1* transcript levels also with CVD [40]. In platelets from healthy donors *F13A1* transcripts correlated with total *RUNX1* and *RUNX1B*-specific transcripts [20]. The association of whole blood *F13A1* with CVD events may reflect the platelet-MK *F13A1*, however, because monocytes also contribute, it would be important to determine in future studies the effect of *RUNX1* mutations on monocyte *F13A1* expression. Our other studies show that the antiplatelet drug ticagrelor, which is used in CVD, enhanced platelet *F13A1* expression in healthy volunteers [20] suggesting that drug therapy modulates platelet *F13A1* expression. Moreover, RUNX1 is an aspirin-responsive transcription factor [40]. Together, these studies may reflect the potential relevance of RUNX1 regulation of *F13A1* to cardiovascular disease.

In summary, our studies provide evidence that *F13A1* is a RUNX1 target and is downregulated in platelets-MK in FPDMM/RHD. Platelet- and MK-driven clot contraction is impaired in RHD, associated with alterations in multiple cellular mechanisms that regulate clot contraction. These findings are also particularly notable because they demonstrate regulation of a coagulation protein – FXIII-A - by a hematopoietic transcription factor, RUNX1, via its effects on MK.

## Data Availability

All data produced in the present work are contained in the manuscript.

## Acknowledgements

We are grateful to the patients who have supported our studies over many years. This study was supported by research funding from NIH (NHLBI), R01 HL137376 and R01 HL109568 to AKR, and R01 HL137207 and HL159006 to LEG and R35 HL150698 to MP, American Heart Association Transformational Project Award 20TPA35490278 to LEG, and a RUNX1 Research Program/Alex’s Lemonade Foundation grant to MP. We thank David E. Ambrose and Amir Yarmahmoodi for discussions and assistance in the flow cytometry studies.

## Author Contributions

FDC, NS and LG performed the research, analysed the data, and contributed to the manuscript; GFM performed the research and analysed the data; LEG and JW designed and performed the immunofluorescence studies; MPL characterized the genetic defects in 2 patients and provided the blood samples; KL and MP generously provided the MK differentiated from the human CD34^+^ cells with RUNX1 knockdown and their expertise and review of the manuscript; AKR conceived, designed and directed the project, participated in all aspects of the research and wrote the manuscript. All authors contributed to the manuscript, have read and approved the manuscript.

