## Supplemental for "Transcription Factor RUNX1 Regulates Coagulation Factor XIII-A (*F13A1*): Decreased Platelet-Megakaryocyte *F13A1* Expression and Clot Contraction in *RUNX1* Haplodeficiency": Supplemental Materials_Carpio-Cano_120524.pdf

### Reagents

All chemicals, including phorbol 12-myristate 13-acetate (PMA), RIPA lysis buffer (25 mM Tris-HCl, pH 7.6, 150 mM NaCl, 1% NP40, 1% sodium deoxycholate, and 0.1% sodium dodecyl sulfate (SDS)) and glass siliconizing reagent Sigmacote were purchased from Sigma-Aldrich (St Louis, MO) and Thermo Fisher Scientific Inc (Waltham, MA). Human Fibronectin (FC010) was from EMD Millipore (Burlington, MA); Trizol Reagent and SuperScript II cDNA synthesis kit was from Invitrogen (Carlsbad, CA); Nuclear and cytoplasmic protein extraction reagent NE-PER from Thermo Fisher Scientific Inc (Waltham, MA). The PCR enzymes, restriction endonucleases and modifying enzymes, luciferase reporter vectors pGL3-Basic and pRL-TK containing the *Renilla* luciferase gene, and the Dual Luciferase Assay kit were from Promega (Madison, WI) and New England Biolabs (Beverly, MA). PCR primers and IRDye 700 probes for electrophoretic mobility shift assays (EMSAs) were synthesized by Integrated DNA Technologies (Coralville, IA). For ChIP analysis, ChIP-It kit was from Active Motif (Carlsbad, CA). Tris-borate EDTA native acrylamide gels were from Bio-Rad Laboratories Inc. (Hercules, CA). Genefect transfection reagent for Luciferase reporter assay was from Molecular Research Laboratories (Sterling, VA). Overexpression vectors pCMV6-XL4 and RUNX1-pCMV6-XL4 were from OriGene Technologies, Inc. (Rockville, MD). The DNA samples were sequenced at GENEWIZ (South Plainfield, NJ). For siRNA knockdown studies, RUNX1 siRNA pools (sc-37677), F13A1 (sc-72083) and control siRNA (sc-44233) were from Santa Cruz Biotechnology, Inc. (Dallas, TX) with Lipofectamine 2000 reagent was from Invitrogen (Carlsbad, CA).

Tissue Factor, human, re-lipidated (4500 L/B) was from American Diagnostic (Windsor, NS Canada), human fibrinogen- Research grade (HCI-0150R) from Haematologic Technology (Essex Junction, VT), Thrombin was from human plasma (T6884), ADP (A2754-1G). Other reagents were obtained as follows: calcium Ionophore A23187 (C9275-1MG), inhibitors Blebbistatin (B0560-1MG) and eptifibatide acetate (SML-1042-10MG) were from Sigma-Aldrich (St Louis, MO); T101(RN2.5) was from Zedira GmbH (Darmstadt, Germany), SFLLRN (PAR1 Agonist peptide 776672-1) was from GenScript (Piscataway, NJ); collagen (#385) from CHRONO-PAR and CHRONO-LUME reagent. Sphero Ultra Rainbow Fluorescent particles (URFP01-30-10K) were from Spherotech Inc. (Lake Forest, IL).

### Methods

#### Quantitative Real-time PCR

Total RNA was isolated from HEL cells using Trizol reagent. 1µg of RNA was reverse transcribed using SuperScript II Reverse Transcriptase. RT- products were amplified with primers (0.1 µM each) for *F13A1* and *GAPDH* shown in Table S3. PCR products were resolved on a 3% agarose gel with ethidium bromide staining.

#### Chromatin Immunoprecipitation (CHIP) assay

HEL cells treated with PMA (10 nM) for 24 h were cross-linked with 1% formaldehyde and sheared enzymatically for 15 min. ChIP analysis was performed with the ChIP-It kit. Chromatin was immunoprecipitated with control IgG and RUNX1 antibody (sc-8564x). *F13A1* promoter regions were amplified by PCR from immunoprecipitated and input samples, with specific primers listed in Table S4. GAPDH was simultaneously amplified. The amplified PCR products were analyzed by gel electrophoresis on 3% agarose gels.

#### Electrophoretic Mobility Shift Assays (EMSA)

Nuclear protein from PMA-treated (10 nM)  HEL cells were extracted with NE-PER Nuclear and Cytoplasmic Extraction Reagents. IRDye-labeled oligonucleotides were double stranded. Nuclear protein (10 µg) was incubated with 50 nM probe and 1 µg Poly(deoxyinosinicdeoxycytidylic) for 20 minutes at RT in a 20-μL reaction, containing 10 mM Tris, 50 mM KCl, 3.5 mM DTT, 0.25% Tween 20, 2.5% glycerol, and 10 mM EDTA before analysis on polyacrylamide gels. For supershift assays, 1 μg RUNX1 antibody (N-20 sc-8563) or control IgG (sc-2025) were incubated with the binding reaction for 20 minutes at RT before the addition of IRDye-labeled probe. Competition assays were performed with 100-fold excess unlabeled probe in 2 μL of nuclear extract and incubated for 10 minutes on ice. The oligonucleotide sequences used to prepare DNA probes are shown Table S5.

**Promoter and plasmid constructs**

Promoter constructs used in transient transfections were generated using promoterless firefly-luciferase reporter gene vector PGL3-Basic (Promega Corp., Madison, WI). Reporter plasmids with a common 3′ end were generated either by unique restriction enzyme digestions or by PCR-based directional cloning using primers as shown in Table S6. Mutant plasmids with deleted or substituted nucleotides in the RUNX1 consensus sites were generated by PCR mutagenesis. The products were verified by sequencing on the ABI Prism 377 (Applied Biosystems, Foster City, CA).

**Luciferase reporter assay**

PMA-HEL cells ($2×10^6$cells) were co-transfected with *F13A1*-promoter-PGL3-luciferase vector (5 μg) and a control vector (promoterless-PGL3-basic vector), pRL-TK (0.1 μg), containing Renilla luciferase gene (Promega) at a ratio of 50:1, using Genefect (Molecular Research Laboratories, Sterling, VA) transfection reagent. After 24 h, cells were lysed, and activity was determined using the Dual-Luciferase Assay System (Promega). Promoter activity was expressed as firefly luciferase activity/Renilla luciferase activity relative to the control PGL3-basic vector. All transfection experiments were performed 3-4 times in triplicate.

**RUNX1 overexpression**

HEL cells ($1.5\times10^6$) were co-transfected with 2 μg each of RUNX1-pCMV6-XL4 or empty vector pCMV6-XL4 (OriGene Technologies, Inc. Rockville, MD) together with 500 ng *F13A1*-promoter-PGL3-luciferase vector using Lipofectamine 2000 and incubated at 37°C with 5% $CO_2$. Luciferase activity and immunoblotting were assessed at 48h.

**RUNX1 and F13A1 downregulation**

*RUNX1* and *F13A1* knockdown were done using *RUNX1* siRNA pools (sc-37677), *F13A1* (sc-72083) and control siRNA (sc-44233) (Santa Cruz Biotechnology Inc, Dallas, TX). HEL cells ($1.5\times10^6$) were co-transfected with *F13A1*-promoter-PGL3-Luciferase vector (500 ng) and RUNX1 siRNA (200 nM, added twice, 48 h apart) or siControl, transferred to 2X medium containing PMA (30 nM), and incubated at 37°C with 5% $CO_2$. Plasmid pRL-TK containing the *Renilla* luciferase gene was used as an internal control. Luciferase activity and protein levels were determined at 96 h.

**Supplemental Table 1. List of antibodies used for immunoblots, ChIP and EMSA studies.**

| Primary Antibody | Source | Reactivity | Supplier | Reference |
|---|---|---|---|---|
| **FXIII-A (B-8)** | Monoclonal Mouse | Human, mouse | Santa Cruz Biotechnology | sc-376312 |
| **FXIII-A (A-subunit)** | Polyclonal Sheep | Human, mouse | Affinity Biologicals | SAF13A-AP |
| **IgG** | Monoclonal Mouse | Human, mouse | Santa Cruz Biotechnology | sc-2025 |
| **MYL9/MYL12A/B (D-9)** | Monoclonal Mouse | Human, mouse | Santa Cruz Biotechnology | sc-48414 |
| **RUNX1 N-20** | Polyclonal Goat | Human, mouse | Santa Cruz Biotechnology | sc-8563 |
| **Phospho-Myosin Light Chain 2 (Thr18/Ser19)** | Polyclonal Rabbit | Human, mouse | Cell Signaling | 3674S |
| **RUNX1** | Polyclonal Goat | Human, mouse | Santa Cruz Biotechnology | sc-8564 |
| **RUNX1** | Monoclonal Mouse | Human, rat, mouse | Santa Cruz Biotechnology | sc-365644 |
| **β-actin** | Polyclonal Mouse | Human, mouse, rat | Santa Cruz Biotechnology | sc-47778 |

**Supplemental Table 2. List of fluorescence-labeled conjugated antibodies.**

| PROTEIN | CONJUGATES | SUPPLIER | REFERENCE |
|---|---|---|---|
| **FXIII-A (EP3372)** | Alexa Fluor-647 | Abcam | ab225018 |
| **IRDye- IgG labeled** | 680/800 RD | Li-Cor Biosciences | 926-68072 |
| **PAC1-FITC** | Fluorescein isothiocyanate | BD Biosciences | 340507 |
| **CD41-FITC** | Fluorescein isothiocyanate | BD Biosciences | 340929 |

**Supplemental Table 3. Oligonucleotide sequences used in Real time reverse transcription - PCR**

| Primers | Strand | Nucleotide Sequence |
|---|---|---|
| ***F13A1*** | F | 5’-GACCTCTCTGGAAGAGGGAA-3’ |
| | R | 5’-GGCATAGATATTGTCCCAGGAT-3’ |
| ***GAPDH*** | F | 5’-GTCTCCTCTGACTTCAACAGCG-3’ |
| | R | 5’-CCTGTTCATTAGATCAGTGGGT-3’ |

**Supplemental Table 4. Oligonucleotide primer sequences used in PCR for ChIP assay.**

| Primers | Position | Strand | Nucleotide Sequence |
|---|---|---|---|
| ***F13A1-Site1*** | -157 to -138 nt | F | 5'-GGGATAACAGGCCAGATGAA-3' |
| ***F13A1-Site1*** | +19 to -1 nt | R | 5'-GACTTCCTCAAACGGACTCG-3' |
| ***F13A1-Site2*** | -280 to 261 nt | F | 5'-CCTGGTACTCCCAGCAACTG-3' |
| ***F13A1-Site2*** | -111 to -130 nt | R | 5'-GGGGAGCTCAGAGGATTTTC-3' |
| ***F13A1-Site3*** | -350 to -332 nt | F | 5'-AAGGAAACCCTCCCAGACC-3' |
| ***F13A1-Site3*** | -266 to -284 nt | R | 5'-GCTGGGAGTACCAGGCAAG-3' |
| ***F13A1-Site4*** | -515 to -496 nt | F | 5'-TGATTCAGGAATGCAGACCA-3' |
| ***F13A1-Site4*** | -373 to -392 nt | R | 5'-GCAGGTCAGCTGCTACTGTG-3' |
| ***F13A1-Site5&6&7*** | -559 to -540 nt | F | 5'-CCTGCAGGAAGTCTTGTGGT-3' |
| ***F13A1-Site5&6&7*** | -551 to -570 nt | R | 5'-ATGCATGCAGTGAGAGCAAC-3' |

### Supplemental Table 5. Oligonucleotide sequences used in DNA probes for EMSA

| Primers | Position | Strand | Nucleotide Sequence |
|---|---|---|---|
| ***F13A1*-Site1** | -115 to -88 nt | F | 5'-CCCTACTGGCTCCAGCTGTGGAGAAGGG-3' |
| ***F13A1*-Site4** | -407 to -378 nt | F | 5'-CCTCCTGCCAAGCCACAGTAGCAGCTGACC-3' |
| ***F13A1*-Site5&6&7** | -553 to -517 nt | F | 5'-GAAGTCTTGTGGTTAGGCGGTGGCTGTGGCTCTGGA-3' |

### Supplemental Table 6. Oligonucleotide sequences used on Luciferase reporter studies

| Primers | Strand | Nucleotide Sequence |
|---|---|---|
| ***F13A1*-Mutant-1** | F | 5'-CCTACTGGCTCCAGCTATGAAAGA-3' |
| | R | 5'-CCCTTCTTCATAGCTGGAGCCAGTAGGGGAGCTCAGAGG-3' |
| ***F13A1*-Mutant-2** | F | 5'-GTACTCCCAGCAACTGGTTACGATGGGGAGGGCAGATC-3' |
| | R | 5'-CTCCCCATCGTAACCAGTTGCTGGGAGTACCAGGCAAG-3' |
| ***F13A1*-Mutant-3** | F | 5'-CCAGACCCTCTGATCATAGGGGACGGGTGG-3' |
| | R | 5'-CCCCTATGATCAGAGGGTCTGGGAGGGTTTCC-3' |
| ***F13A1*-Mutant-4** | F | 5'-CCAAGACAAAGTAGCAGCTGACCTGCCAGGC-3' |
| | R | 5'-GGTCAGCTGCTACTTTGTCTTGGCAGGAGGG-3' |
| ***F13A1*-Mutant-5** | F | 5'-GTCTTATGATTAGACGATGGCTATGACTCTGGGATGATTC-3' |
| | R | 5'-CAGAGTCATAGCCATCGTCTAATCATAAGACTTCCTGCAG-3' |
| ***F13A1*-Mutant-6** | F | 5-'GGTTAGGCGGTGGCTATGACTCTGGGATGATTCAGG-3' |
| | R | 5'-CCCAGAGTCATAGCCACCGCCTAACCACAAGACTTCC-3' |
| ***F13A1*-Mutant-7** | F | 5'-GTCTTGTGGTTAGACGATGGCTGTGGCTCTGGGATG-3' |
| | R | 5-'CAGCCATCGTCTAACCACAAGACTTCCTGCAGGAATGG-3' |
